# Sex differences in brain development in fetuses and infants who are at low or high likelihood for autism

**DOI:** 10.1101/2021.03.08.21251862

**Authors:** L. M. Villa, S. Hampton, E. Aydin, R. Tait, M. J. Leming, A. Tsompanidis, I. Patterson, C. Allison, T. Austin, J. Suckling, S. Baron-Cohen, R. J. Holt

## Abstract

**Background:** It is unknown whether the neural underpinnings of autism are present *in utero*. In addition, it is unclear whether typical neural sexual differentiation, which is associated with the development of autism, is evident *in utero*. We longitudinally investigated fetal and infant sex differences in brain structure and function, and differences in brain development in those at low and high likelihood for autism. Here, we use the term ‘typical’ interchangeably with the term ‘low-autism likelihood’.

**Methods:** Participants were longitudinally studied *in utero* first at 30-33 weeks of gestation, and then as infants 8-12 weeks after birth. We compared total brain volumes and resting-state functional connectivity between 15 female and 15 male low-autism likelihood fetuses (defined as having no first degree autistic relative). We also compared the brain structure and function of these 30 fetuses to a rare group of 11 fetuses (5 females and 6 males) who had an autistic mother or sibling, and therefore a higher likelihood of developing autism. Although a small sample, the high-autism likelihood group are reported as they are challenging to recruit. Additionally, we correlated sex differences in functional connectivity with autism likelihood group differences across the fetal and infant brains.

**Results:** There was a group-by-sex interaction in fetal total brain volume. Typical males, on average, showed faster total brain volume growth in the perinatal period than typical females. The high-autism likelihood group showed lower resting-state functional connectivity at both time-points compared to the typical group, and regions indicating sex differences overlapped with those associated with high-autism likelihood group differences in functional connectivity.

**Conclusions:** *In utero* sexual differentiation of brain structure was more pronounced in fetuses with a high likelihood for autism. Moreover, sexual differentiation of the fetal and infant brain may overlap with the neural development of autism.

## 1. Introduction

Males are more likely to be diagnosed with autism than females, with the male to female ratio in autism diagnoses being around 3:1 (Baron-Cohen et al., 2011; Loomes, Hull, & Mandy, 2017). This has led to speculation about whether biological differences between males and females are partly responsible for this gender imbalance, with several studies identifying a relationship between sexual differentiation of the brain and the development of autism (Auyeung et al., 2012; Baron-Cohen, Knickmeyer, & Belmonte, 2005; Beacher et al., 2012; Floris, Lai, Nath, Milham, & Di Martino, 2018; Lai et al., 2013).

The neural underpinnings of autism appear to arise early in life. Lower functional connectivity has been found in autistic children between the ages of 2-4 years (Dinstein et al., 2011; Shen et al., 2016) and larger regional and total brain volumes in autistic children appear to be present by the age of two (Conti et al., 2017; Conti et al., 2016, 2015; Fingher et al., 2017; Hazlett et al., 2011; Retico et al., 2016; Wolff, Jacob, & Elison, 2018; Xiao et al., 2014), with greater cortical surface area being present in autistic children potentially as early as six months after birth (Hazlett et al., 2017).

Moreover, studies investigating typical sexual differentiation suggest that sex differences in both brain structure and function also develop early in life and are present by birth (Benavides et al., 2019; Dean et al., 2018; Gao, Alcauter, Smith, Gilmore, & Lin, 2015; Gilmore et al., 2007; Gilmore et al., 2012; Holland et al., 2014; Knickmeyer et al., 2008, 2014, 2017; Salzwedel et al., 2019; Shi et al., 2011; Tanaka, Matsui, Uematsu, Noguchi, & Miyawaki, 2013; Uematsu et al., 2012), with the most consistent finding being that males on average show larger total brain volumes and faster brain volume growth than females (Benavides et al., 2019; Dean et al., 2018; Gilmore et al., 2007; Holland et al., 2014; Knickmeyer et al., 2008, 2014, 2017; Shi et al., 2011; Tanaka et al., 2013; Uematsu et al., 2012).

However, it remains unclear whether both the neurobiological origins of autism and typical sexual differentiation of the brain occur prenatally and whether they are related. Currently, no study has investigated prenatal differences in brain development related to autism, and it is still unclear whether sex differences within the brain are present prenatally, as some studies have reported subtle structural and functional prenatal sex differences (Andescavage, Duplessis, et al., 2017; Conte et al., 2018; Kyriakopoulou et al., 2017; Wheelock et al., 2019) whilst others have been unable to detect any (Andescavage, Du Plessis, et al., 2017; Dubois et al., 2010; Scott et al., 2011).

To address these questions, we tested for sex differences longitudinally in fetal and infant brain development, as well as testing for differences between low- and high-autism likelihood. The term ‘high-autism likelihood’ refers to elevated genetic likelihood for autism as this group were selected based on having a first degree relative with an autism diagnosis, making their likelihood for autism around 20-30% (Lauritsen, Pedersen, & Mortensen, 2005; Ozonoff et al., 2011; Werling & Geschwind, 2015). This represents a hard-to-recruit group of pregnancies since women whose fetus is at high-autism likelihood – around 1-2% of the population (Baird et al., 2006; Maenner et al., 2020; Sun et al., 2019) – need to be willing to travel for research and consent for repeat MRI in the specified time period during pregnancy.

Participants were assessed longitudinally at two time-points, first at 30-33 weeks of gestation and later at 8-12 weeks after birth. To investigate typical sexual differentiation *in utero* and during early infancy, we compared the total brain volumes and resting-state functional connectivity of typical males and females. We use the term ‘typical’ and ‘low-autism likelihood’ interchangeably, whilst acknowledging that in the typical group it is not possible to say with any certainty that none of these fetuses or infants will not go on to develop autism. The term ‘typical’ in this context refers to the group having a typical autism likelihood of 1-2%, based on autism prevalence (Baird et al., 2006; Maenner et al., 2020; Sun et al., 2019). We further acknowledge that as they did not have a diagnosis of autism, those in the ‘high-autism likelihood’ group may also be typically developing.

We hypothesized that typical male infants would on average have larger total brain volumes than typical female infants, based on previous reports (Andescavage, et al., 2017; Benavides et al., 2019; Dean et al., 2018; Dubois et al., 2010; Gilmore et al., 2007; Knickmeyer et al., 2008, 2014, 2017; Scott et al., 2011; Shi et al., 2011; Uematsu et al., 2012). As past research has found both structural and functional neonatal sexual differentiation of the cerebellum, hippocampus, and amygdala (Dean et al., 2018; Gilmore et al., 2007; Knickmeyer et al., 2014, 2017; Salzwedel et al., 2019; Uematsu et al., 2012; Wolff et al., 2018), we also hypothesized that the resting-state functional connectivity of these regions would show sexual differentiation in typical infants.

We also compared the total brain volumes and resting-state functional connectivity of typical fetuses and infants to those with a high likelihood for developing autism, defined by having an autistic mother or autistic sibling. We hypothesized that high-autism likelihood participants would show greater total brain volumes at both the fetal and infant time-points than typical participants, based on results in older children (Hazlett et al., 2017; Hazlett et al., 2011; Shen et al., 2013; Wolff et al., 2018). We further hypothesized that the high-autism likelihood participants would show differences in resting-state functional connectivity. Finally, to study the relationship between sex differences and autism-related differences in brain development, we investigated whether brain regions showing sexual differentiation in brain function colocate with high-autism likelihood-related differences.

## 2. Methods

### 2.1. Participants

Participants in the typical group were recruited from the Rosie Maternity Hospital, Cambridge, using posters and by speaking to individuals when attending their medical appointments. Identifying mothers whose offspring were at a greater likelihood of developing autism, were in the third trimester of pregnancy, and were able to participate in the study, posed significant challenges due to the commonality of this group, and geographical and timing restrictions. Participants in the high-autism likelihood group were recruited through the Cambridge Autism Research Database (CARD), support groups across the UK, social media, and advertisements in magazine adverts. The typical (low-autism likelihood) group comprised pregnant women whose infants had no self-reported first degree relative with a diagnosed autism spectrum condition. The high-autism likelihood group comprised either pregnant women with a diagnosis of autism or pregnant women who already had a child with a diagnosis of autism. Only mothers with a singleton pregnancy were eligible to take part, and no participants reported smoking or regularly consuming alcohol during pregnancy. Favorable ethical opinion was obtained (REC reference number: 12/EE/0393) and all mothers gave their written informed consent.

Participants were assessed with both structural and functional MRI scans: one *in utero* fetal MRI scan during 30-33 weeks of gestation, and one infant MRI scan at 8-12 weeks after birth. Infant scans were conducted during natural sleep with no sedation. Thirty-two (32) typical and 11 high-autism likelihood participants were enrolled. Structural data from two typical participants were excluded due to motion artefacts, resulting in 30 typical and 11 high-autism likelihood fetuses with useable structural data. Functional data from a further four typical participants were also excluded due to motion artefacts, resulting in 26 typical and 11 high-autism likelihood fetuses with useable functional data.

At the second (infant) time-point, one high-autism likelihood and two typical infants were unable to be followed-up. Therefore, 30 typical and 10 high-autism likelihood infants underwent scanning. Structural data from one high-autism likelihood and 10 typical infants were excluded due to the infant’s inability to achieve sleep in the scanner. Further data from two high-autism likelihood infants were excluded, one due to movement artefacts, and one due to a technical failure. This resulted in 20 typical and 7 high-autism likelihood infants with useable structural data at both time-points. Functional data from a further four typical infants were excluded due to movement artefacts, resulting in 16 typical and 7 high-autism likelihood infants with useable functional data at both time-points.

### 2.2. MRI acquisition

All MR images were obtained at the Rosie Hospital, University of Cambridge, UK, using a 1.5T MR450w Artist, GE Healthcare scanner. Structural images were acquired using the balanced steady-state gradient echo sequence, Fast Imaging Employing Steady-state Acquisition (FIESTA) with the following parameters: repetition time (TR) = 3.5ms, echo time (TE) = 1.4ms, flip angle (FA) = 55°, 172 slices with voxel size of 1.875×1.875×1mm^3^, and a field of view of 256×256mm^2^ voxels. BOLD sensitive echo-planar images were acquired using the following parameters: TR = 2000ms, TE = 40ms, FA = 90°, 150 volumes, 3150 slices of 4mm thickness, voxel size of 5.625×5.625×4mm^3^, field of view of 64×64mm^2^ voxels, and a scan time of 5.00 minutes.

### 2.3. Structural image pre-processing

Fetal brain images were manually reoriented using the Reorient tool in ITK-SNAP (Yushkevich et al., 2006) as *in utero* head movement led to mismatches between image orientation codes and the actual orientation of fetal brains. This step was not necessary for infant structural images as infants were scanned during natural sleep with cushioned head padding to minimize movement within the scanner. The spatial origin was set to the anterior commissure fiber tract, permitting accurate co-registration of reoriented images. All images were manually skull-stripped with the resulting ROI extending to the skull/cerebrospinal fluid interface and to the base of the cerebellum.

Using a sex balanced random sample of participants, with both fetal and infant images, a study-specific structural template was generated using Advanced Normalization Tools (ANTs; Avants, Tustison, & Song, 2009). The STA31 template, part of a spatiotemporal MRI atlas for segmentation of early brain development (Gholipour et al., 2017), together with all fetal and infant structural images were registered to the study-specific template using ANTs (Avants et al., 2009). To avoid interpolation artefacts, STA31-to-study-specific template and subject-to-study-specific template spatial mappings were composed into a single inversely symmetrical STA31-to-subject transformation.

As it was not possible to directly segment brain images into grey and white matter due to the absence of grey/white matter contrast during this developmental period, STA31 parcellations, which include parcellations of grey matter, white matter, and cerebrospinal fluid, were re-sampled into acquisition space to indirectly estimate grey matter properties within the brain. The sum of combined re-sampled STA31 parcellations was then used to estimate total brain volume from the structural images.

### 2.4. Functional image pre-processing

Fetal functional images were manually reoriented and had their spatial origin set, as previously described for structural images. A whole brain ROI was manually drawn on a reference (i.e. first) frame of each fetal and infant functional dataset. A rigid-body functional reference frame-to-structural image transformation was then generated using ANTs (Avants et al., 2009). Functional-to-structural and structural-to-study-specific template spatial mappings were composed into a single functional-to-study-specific template transformation.

Due to *in utero* head movement, sub-blocks of frames in approximate alignment were manually identified, averaged and co-registered to the reference frame. Once aligned with the reference frame, functional images were skull-stripped and pre-processed using the speedypp toolkit (Patel et al., 2014). Pre-processing consisted of slice-timing correction, rigid-body frame alignment, intensity normalization, followed by wavelet despiking and confound signal regression. Applying the functional-to-study-specific template transformation with a half resolution field of view resulted in all pre-processed functional images being re-sampled into functional template space. Although they had less movement than the fetal images, the infant functional images were pre-processed using the same steps to mitigate against bias.

### 2.5. Statistical analyses

#### 2.5.1. Total brain volume analyses

To investigate sex differences in brain structure in the typical group at each time-point separately, an ANCOVA was used with total brain volume as the dependent variable and sex as the independent variable. For fetal analyses, gestational age was included as a covariate, whereas for infant analyses, age since conception and birth weight were used as covariates. To investigate differences between typical vs high-autism likelihood groups at each time-point separately, an ANCOVA was used with total brain volume as the dependent variable and group and sex as the independent variables. A group-by-sex interaction was also included. For fetal analyses, gestational age was used as a covariate. For infant analyses, age since conception, and birth weight were used as covariates. To investigate differences in total brain volume growth, a repeated measures ANCOVA was used to investigate a sex-by-time interaction within the typical group, with total brain volume as the dependent variable, and sex and time as the independent variables. A group-by-time interaction was also conducted, with total brain volume as the dependent variable, and group and time as the dependent variables, using sex as a covariate.

#### 2.5.2. Seed-based resting-state functional connectivity analyses

Based on previous findings (Dean et al., 2018; Gilmore et al., 2007; Knickmeyer et al., 2014, 2017; Salzwedel et al., 2019; Uematsu et al., 2012; Wolff et al., 2018), the left and right amygdala, left and right hippocampus, and left and right cerebellum were used as seed regions. Seed regions, located using the STA31 template (Gholipour et al., 2017), were used to investigate both typical sex differences and group differences in resting-state functional connectivity. Each seed’s mean time-series was regressed on the time-series of all other intracerebral voxels using FSL FEAT (Woolrich, Ripley, Brady, & Smith, 2001). Subsequently, statistical tests were conducted at each voxel across the whole brain, excluding the respective seed region, with permutation-based methods applied for all statistical inferences, via FSL Randomise (Jenkinson, Beckmann, Behrens, Woolrich, & Smith, 2012). Each statistical test used 100,000 permutations, threshold-free cluster enhancement, and a Family-wise error (*pcorr* < 0.05) correction for multiple comparisons. Tests to investigate differences over time were conducted by subtracting each participant’s fetal scan data from their infant scan data, and inference conducted on the subtracted images.

To investigate between-sex differences within the typical group, and between-group differences, gestational age was used as a covariate for fetal analyses, and age since conception for infant analyses. At each time-point separately, a group-by-sex interaction was also conducted, with gestational age used as a covariate at the fetal time-point, and age since conception at the infant time-point. To investigate time-related sex and group differences, a sex-by-time interaction within the typical group was conducted, with sex and time as the dependent variables, and a group-by-time interaction was conducted, with group and time as the dependent variables and sex as a covariate.

#### 2.5.3. Relationship between sex differences and high-autism likelihood differences in resting-state functional connectivity

For each seed, between-sex and between-group differences in resting-state functional connectivity had a respective *F*-score map. We investigated whether brain regions with between-sex differences were colocated with regions of between-group differences. To achieve this, for each seed, we parcellated the corresponding *F*-score map into 108 regions of the brain, based on the STA31 template (Gholipour et al., 2017). For each seed and for each region, we extracted each parcel’s respective between-sex and between-group *F*-scores, log transformed them to achieve approximately normal distributions of scores, and then correlated regional *F*-scores between contrasts. This was done hypothesizing that if a region that showed sex differences were also a region that showed high-autism likelihood differences in resting-state functional connectivity, then there should be a positive correlation between corresponding *F*-scores.

Thus, for each of the six seed regions (left and right amygdala, left and right hippocampus, and left and right cerebellum) there were 107 correlations (number of parcels minus the seed region). Each of these correlations were then Bonferroni corrected (0.05/642). Any significant positive correlations were converted into *r*^*2*^ values, as a measure of effect size. This method, using similar procedures, has been used previously (Villa et al., 2020).

## 3. Results

### 3.1. Demographics

Demographic details of fetuses and infants are shown in **Table 1** and **Table 2**, respectively. There were no significant demographic differences between typical male and typical females, or between typical and high-autism likelihood groups, except with infant birth weights, where high-autism likelihood infants had greater average birth weights, *t*(25) = 2.15, *p* = 0.042, *d* = 0.94. Full details of the sample and the statistical tests on these measures are shown in supplementary information (**S1**).

**Table 1.** Demographic details of the fetal groups, with standard deviations in parentheses.

|  | **Mean Gestational Age**<br>**(Days)** |
| --- | --- |
| **Typical males**<br>[Structural],<br>(*n* = 15) | 221.60<br>(9.30) |
| **Typical females**<br>[Structural],<br>(*n* = 15) | 226.80<br>(6.78) |
| **Typical males**<br>[Functional],<br>(*n* = 12) | 227.08<br>(6.65) |
| **Typical females**<br>[Functional],<br>(*n* = 14) | 221.07<br>(9.41) |
| **Typical group**<br>[Structural],<br>(*n* = 30, 50% female) | 224.20<br>(8.42) |
| **High-autism likelihood group**<br>[Structural],<br>(*n* = 11, 45.5% female) | 225.18<br>(10.86) |
| **Typical group**<br>[Functional],<br>(*n* = 26, 53.8% female) | 223.85<br>(8.65) |
| **High-autism likelihood group**<br>[Functional],<br>(*n* = 11, 45.5% female) | 225.18<br>(10.86) |

**Table 2.** Demographic details of the infant groups, with standard deviations in parentheses.

|  | Mean Age Since<br>Conception<br>(Days) | Mean Birth<br>weight<br>(Grams) |
| --- | --- | --- |
| **Typical males**<br>[Structural],<br>(*n* = 10) | 354.00<br>(13.55) | 3564.00<br>(462.59) |
| **Typical females**<br>[Structural],<br>(*n* = 10) | 354.10<br>(13.30) | 3265.50<br>(363.99) |
| **Typical males**<br>[Functional],<br>(*n* = 9) | 353.78<br>(14.35) | 3526.67<br>(474.41) |
| **Typical females**<br>[Functional],<br>(*n* = 7) | 354.57<br>(15.61) | 3265.00<br>(398.56) |
| **Typical group**<br>[Structural],<br>(*n* = 20, 50.0% female) | 354.05<br>(13.07) | 3414.75<br>(433.09) |
| **High-autism likelihood**<br>[Structural],<br>(*n* = 7, 42.9% female) | 353.43<br>(17.68) | 3825.57<br>(445.72) |
| **Typical group**<br>[Functional],<br>(*n* = 16, 43.8% female) | 354.13<br>(14.40) | 3412.19<br>(448.94) |
| **High-autism likelihood**<br>[Functional],<br>(*n* = 7, 42.9% female) | 353.43<br>(17.68) | 3825.57<br>(445.72) |

### 3.2. Imaging analyses

For brevity, only significant results are described in detail. Statistics for non-significant results are shown in supplementary information (**S2**).

#### 3.2.1 Fetal total brain volumes

There were no significant between-sex differences, within the typical group, nor were there significant between-group differences. There was a significant group-by-sex interaction in total brain volume, *F*(1, 36) = 5.04, *p* = 0.031, η_p_ ^2^ = 0.123, whereby males and females within the typical group did not differ in total brain volume, but differences were observed within the high-autism likelihood group, with males in the high-autism likelihood group having larger brain volumes; **Figure 1**.

**Figure 1.**
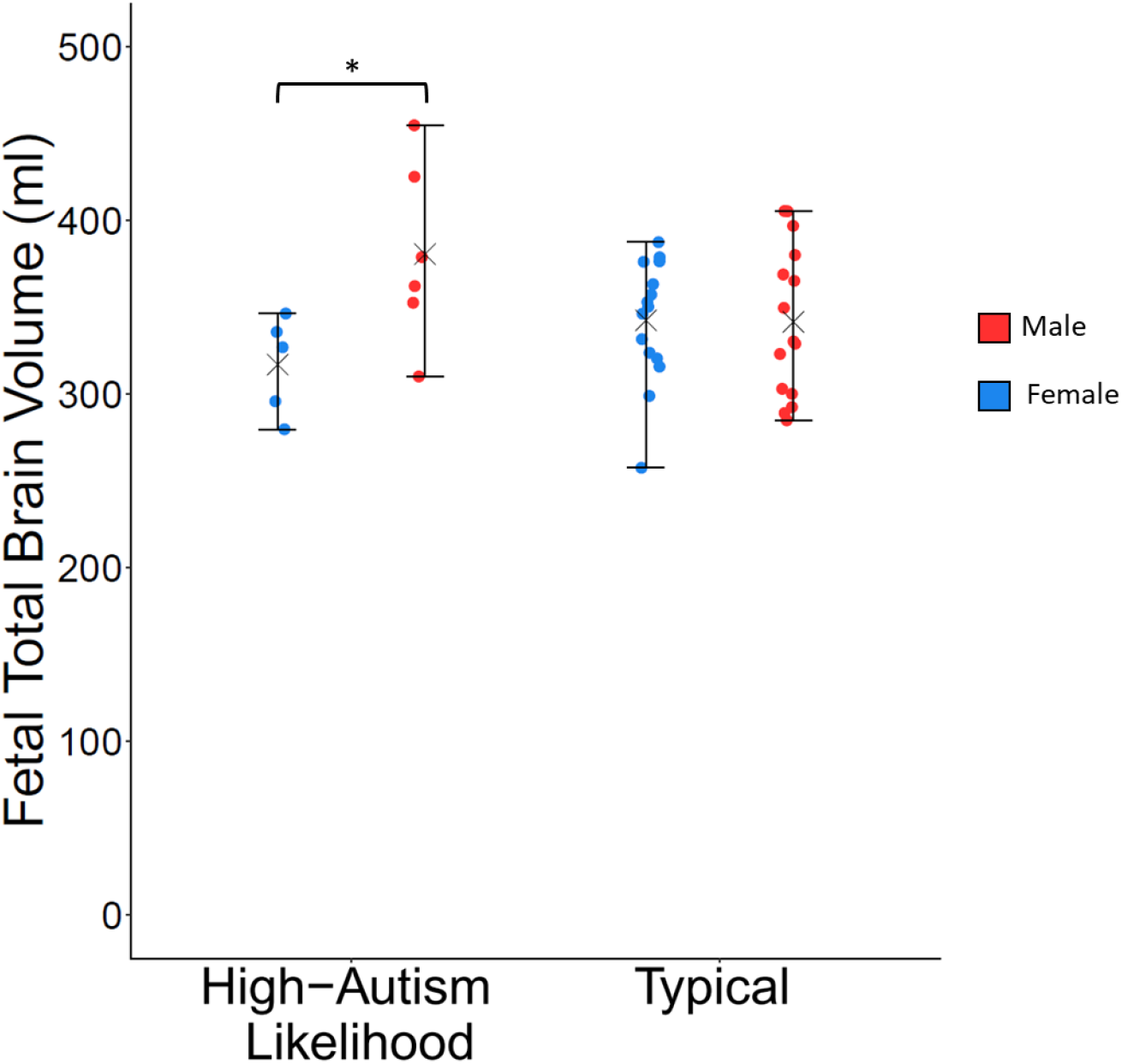
Fetal total brain volumes for males and females in typical and high-autism likelihood groups, where there was a significant group-by-sex interaction.

#### 3.2.2 Fetal brain functional connectivity

There were no significant between-sex differences or group-by-sex interactions. Fetuses within the high-autism likelihood group showed lower resting-state functional connectivity between the left cerebellum seed and the right inferior parietal lobule, right supramarginal gyrus, right supplementary motor area, right medial frontal gyrus, and right midcingulate; **Figure 2** and **Table 3**.

**Table 3.** Details of the significant clusters where the fetal high-autism likelihood group showed lower resting-state functional connectivity.

| Cluster | Number of voxels | Peak $p_{corr}$ value | Peak $t$-score | Peak STA Coordinates | Cohen's $d$ | Regions showing greater connectivity (peak region in bold) |
| --- | --- | --- | --- | --- | --- | --- |
| Cluster 2 | 40 | 0.040 | 4.25 | 54, 59, 53 | 1.49 | **Right inferior parietal lobule**, right supramarginal gyrus |
| Cluster 1 | 18 | 0.047 | 4.15 | 38, 32, 57 | 1.59 | **Right supplementary motor area**, right medial frontal gyrus, right midcingulate |

**Figure 2.**
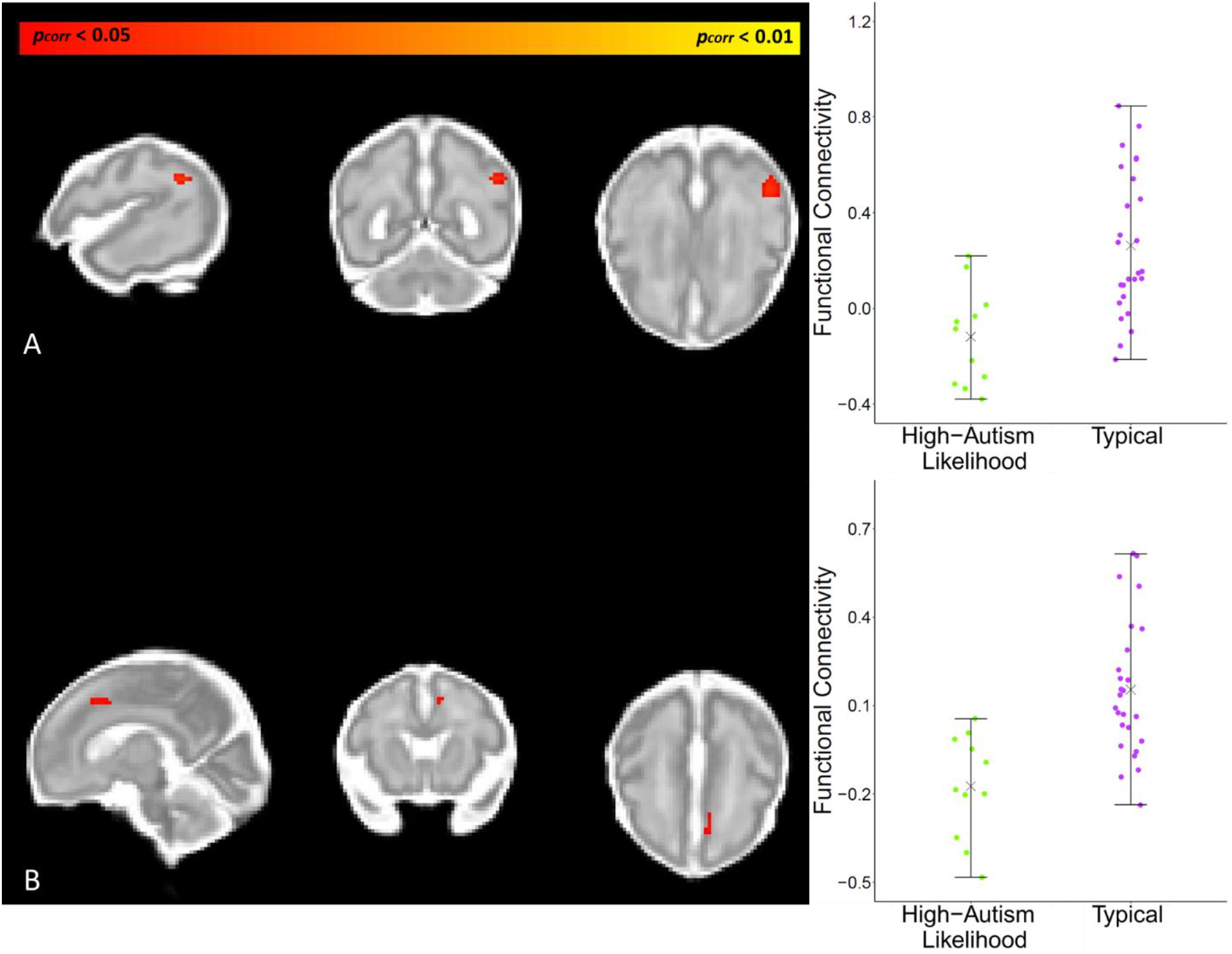
Differences in fetal resting-state functional connectivity between the typical and high-autism likelihood groups. A: Left Cerebellum Seed, Cluster 2 B: Left Cerebellum Seed, Cluster 1.

#### 3.2.3. Infant total brain volumes

There were no significant between-sex differences, within the typical group, nor were there between-group differences in infant total brain volumes, nor was there a significant sex-by-group interaction (see supplementary information (**S2**) for details).

#### 3.2.4 Infant brain functional connectivity

There were no significant between-sex differences, within the typical group, nor were there any significant sex-by-group interactions. Infants within the high-autism likelihood group had lower resting-state functional connectivity between the right amygdala seed and the bilateral precuneus, bilateral posterior cingulum, left calcarine, left cuneus, bilateral superior occipital gyrus, and bilateral middle occipital gyrus; **Figure 3** and **Table 4**.

**Table 4.** Details of the significant clusters where the infant high-autism likelihood group showed lower resting-state functional connectivity.

| Cluster | Number of voxels | Peak *p<sub>corr</sub>* value | Peak *t*-score | Peak STA Coordinates | Cohen's *d* | Regions showing greater connectivity (peak region in bold) |
| --- | --- | --- | --- | --- | --- | --- |
| Cluster 3 | 384 | 0.0063 | 6.15 | 31, 64, 48 | 2.31 | Left posterior cingulum, left calcarine, left cuneus, left superior occipital gyrus, left middle occipital gyrus, **left precuneus** |
| Cluster 2 | 270 | 0.023 | 4.42 | 48, 71, 46 | 1.69 | Right superior occipital gyrus, **right middle occipital gyrus**, right angular gyrus |
| Cluster 1 | 17 | 0.045 | 4.19 | 38, 58, 48 | 2.02 | **Right posterior cingulate**, right precuneus |

**Figure 3.**
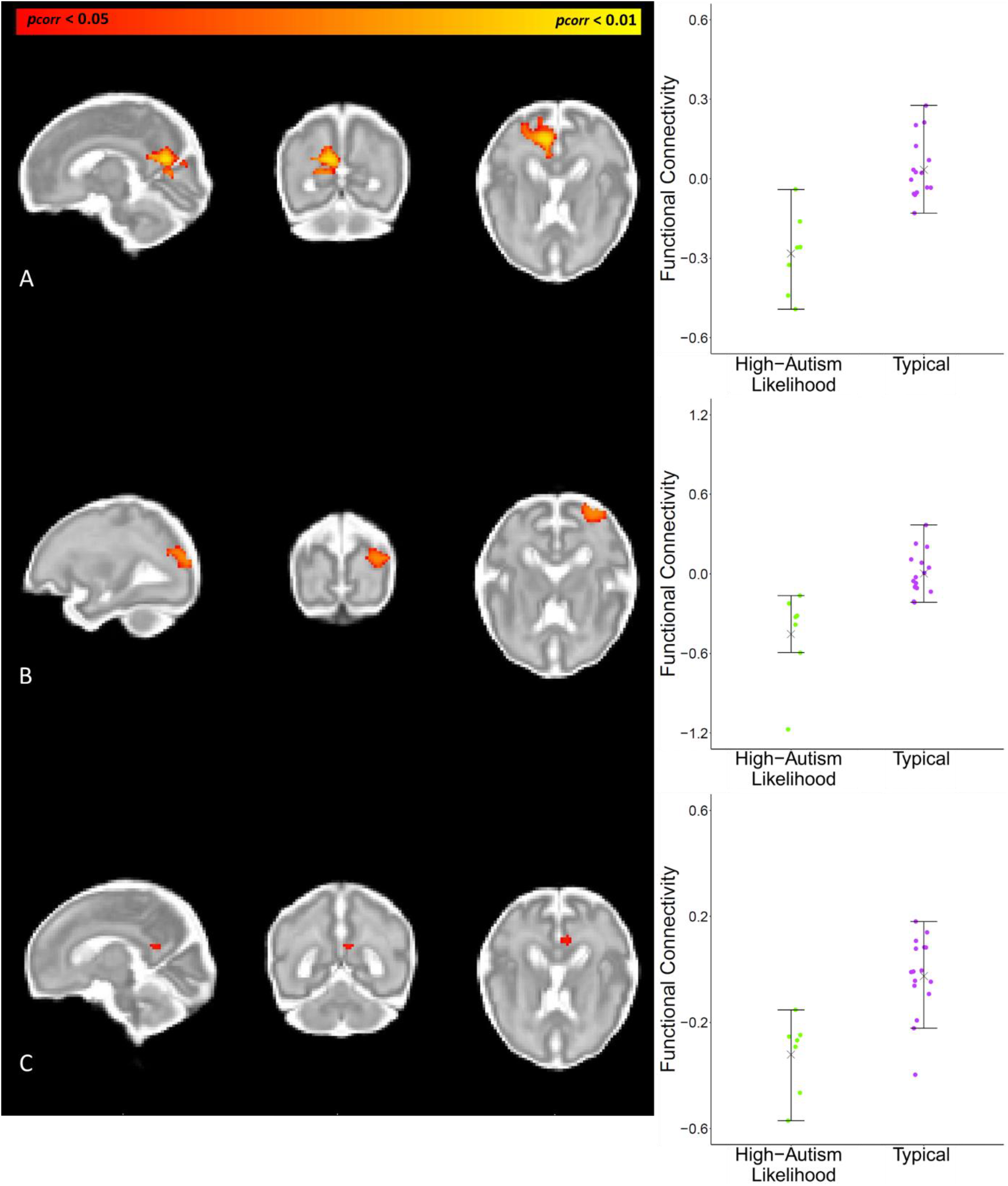
Between-group differences in infant resting-state functional connectivity. A: Right Amygdala Seed, Cluster 3. B: Right Amygdala Seed, Cluster 2. C Right Amygdala Seed, Cluster 1. In all cases, functional connectivity was reduced in the high-autism likelihood group.

#### 3.2.5. Interaction effects in brain volume and functional connectivity with time

There was a significant sex-by-time interaction within the typical group. Typical males showed faster growth in total brain volume than typical females, *F*(1, 16) = 8.84, *p* = 0.009, *η*_*p*_^2^ = 0.27; **Figure 4**. There were no significant group-by-time interactions with either total brain volume or resting-state functional connectivity.

**Figure 4.**
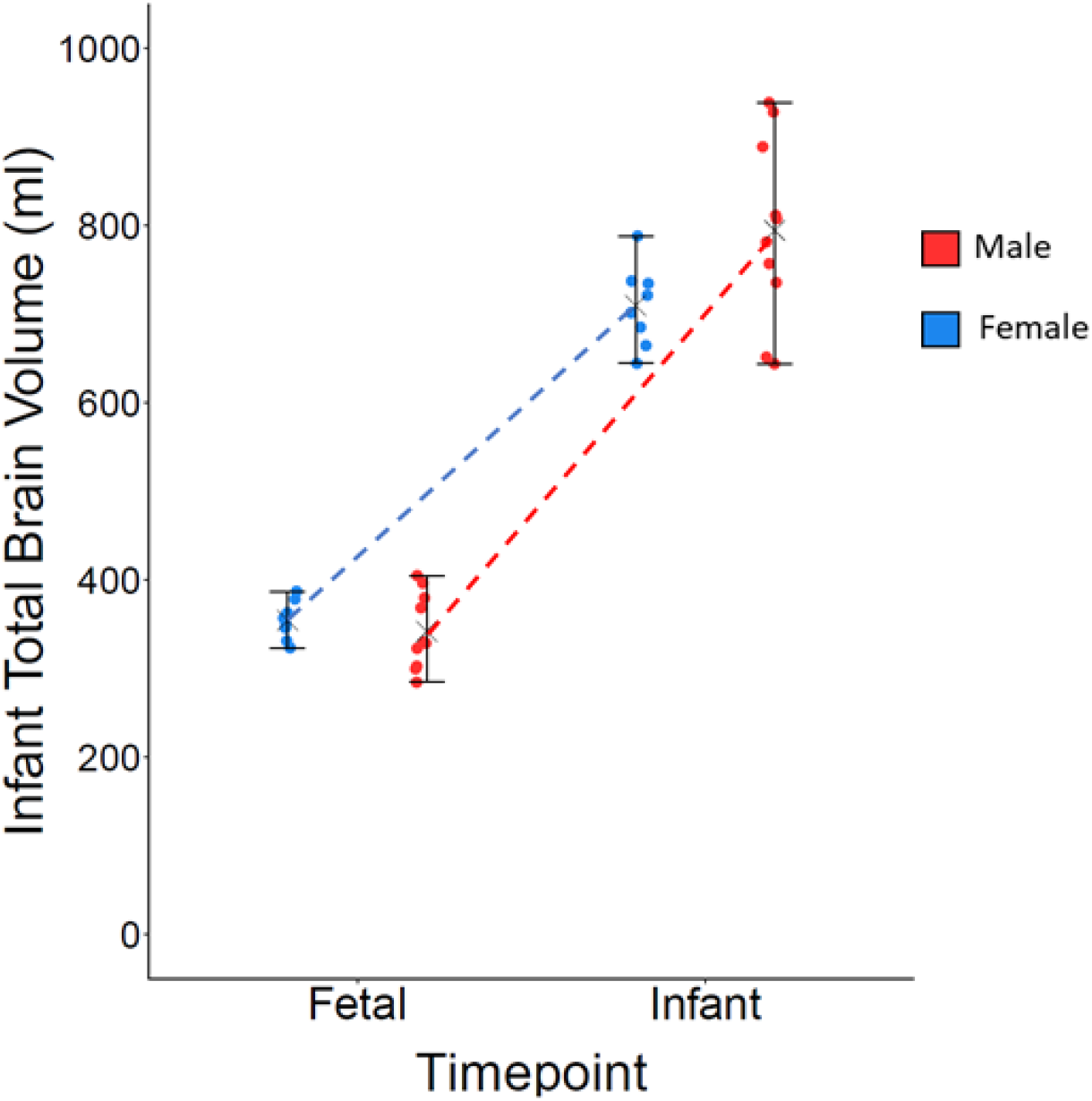
A significant sex-by-time interaction in total brain volume growth, within the typical group.

#### 3.2.6. Relationship between sex differences and high-autism likelihood differences in resting-state functional connectivity

Of the 642 correlations of between-sex and between-group effects in fetal resting-state functional connectivity, 199 survived Bonferroni correction, with 91 (45.72%) being positive correlations. Of the correlations of infant data, 161 survived Bonferroni correction, with 74 (45.96%) being positive. Details of the distribution of both positive and negative correlations are shown in supplementary information (**S3**). Multiple brain regions that showed between-sex effects also showed high-autism likelihood differences at both the fetal and infant time-points. A key effect is shown in **Figure 5** where the *r*^2^ values for the right hippocampus seed at infant and fetal time-points are displayed. Full details for all seeds are shown in supplementary information (**S3**). For fetuses, regions showing the strongest effects across seeds were: left central vermis, right Rolandic operculum, left anterior vermis, left inferior frontal gyrus, left gyrus rectus, right precentral gyrus, left olfactory cortex, right amygdala, right paracentral lobule, and left amygdala. For infants, regions showing the strongest effects across seeds were: right medial orbitofrontal cortex, right superior occipital cortex, left medial orbitofrontal cortex, left superior occipital cortex, left supplementary motor area, right amygdala, and right putamen. **Figure 5**. Effect size, *r*^2^, values across the right hippocampus seed, when correlating sex difference effects with high-autism likelihood differences.

**Figure 5.**
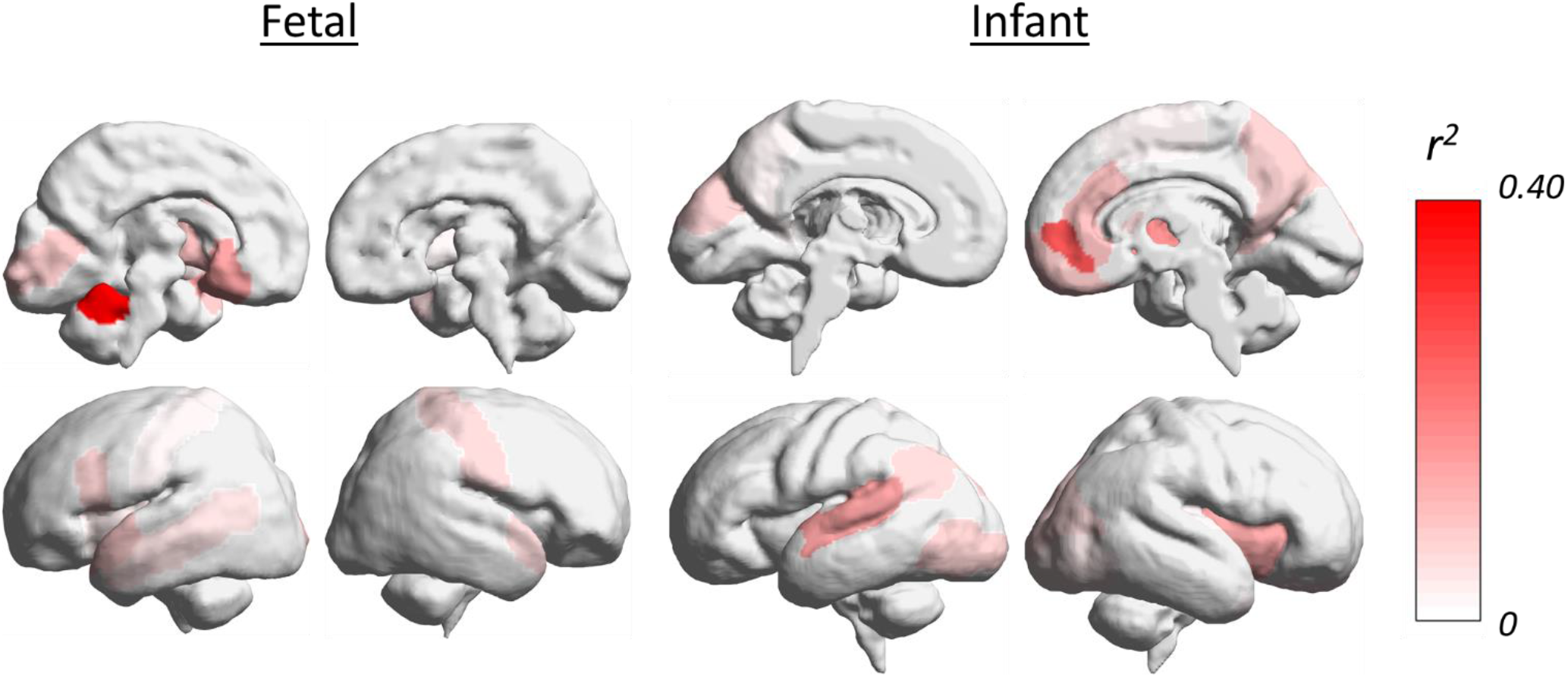
Effect size, r^2^, values across the right hippocampus seed, when correlating sex difference effects with high-autism likelihood differences.

## 4. Discussion

Conducting a longitudinal study attempting to collect *in utero* and infant neuroimaging data including mothers who have autism, or have a child with autism, are pregnant during the third trimester, and are willing and able to participate in the study is extremely challenging. Nevertheless, this is the first longitudinal study to investigate whether a familial history of autism is related to prenatal and early infant neurodevelopment, and the first to investigate longitudinally both prenatal and early infant neurotypical sexual differentiation of both brain structure and function. In the typical group, we found that male total brain volumes on average had a faster rate of growth than those of females. There was also a group-by-sex interaction in fetal total brain volume and group differences in resting-state functional connectivity at both fetal and infant time-points. Moreover, multiple brain regions that showed indications of sex differences in resting-state functional connectivity were also regions that showed autism-related differences in both fetuses and infants.

We did not find any overall sex differences in either brain structure or function at the fetal time-point, which is not surprising given that past studies have also failed to find prenatal sex differences (Andescavage, et al., 2017; Dubois et al., 2010; Scott et al., 2011). The lack of overall sex differences in infant total brain volume does differ from past studies, which have consistently reported neonatal and infant sex differences in total brain volume at birth and within the first year of life (Benavides et al., 2019; Dean et al., 2018; Gilmore et al., 2007; Knickmeyer et al., 2008, 2014, 2017; Shi et al., 2011; Uematsu et al., 2012), but it is likely that our infant analyses were underpowered to detect this sex difference within the typical group, which may account for this discrepancy. However, following longitudinal assessment of both prenatal and postnatal data, we found that typical male total brain volumes, on average, grew at a faster rate than typical females. This result is in line with past research (Holland et al., 2014; Tanaka et al., 2013) and suggests that whilst overall sex differences may not be apparent prenatally, in terms of total brain volume, the process of brain growth begins to differentiate between typical males and females from the 3^rd^ trimester.

There were no overall group differences in total brain volume and thus no evidence for the autism-related brain overgrowth that has been observed in infants as young as 6 months old (Hazlett, et al., 2017), though it should be noted that as none of those in the high-autism likelihood group were clinically diagnosed with autism, this was therefore not a direct test of autism-related brain overgrowth. However, we found a significant group-by-sex interaction in fetal total brain volume, by which male fetuses had significantly larger total brain volumes than females within the high-autism likelihood group. Postnatally, this group-by-sex interaction was no longer present, with there being a trend towards males in both groups having greater total brain volumes than females. These findings may be interpreted as high-autism likelihood males showing earlier sexual differentiation, with typical males possibly showing this differentiation later in development, as a “catch-up” effect, accounting for the lack of a significant group-by-age interaction at the infant time-point. Males have higher levels of sex steroid hormones prenatally, following the activation of the fetal testes, with a surge in their levels between the first and second trimesters (Baron-Cohen, et al., 2004; Baron-Cohen et al., 2011; Hines, Constantinescu, & Spencer, 2015). We have previously reported that sex steroid hormone levels, estradiol in particular, are higher in the amniotic fluid of males later diagnosed with autism than controls (Baron-Cohen et al., 2015, 2020), and correlate with the development of autistic traits (Auyeung et al., 2012, 2009). This greater exposure to sex steroid hormones during gestation may contribute towards the earlier sexual differentiation of total brain volume in high-autism likelihood males. However, this suggestion is speculative and further investigation using a larger cohort, alongside measurements of fetal sex steroid hormones, are required to explore this further.

The high-autism likelihood group showed patterns of functional underconnectivity at both the fetal and infant time-points. At the fetal time-point, the high-autism likelihood group showed underconnectivity between the left cerebellum seed and multiple regions including the medial frontal gyrus, supplementary motor area, midcingulate, and inferior parietal lobule. Furthermore, at the infant time-point, the high-autism likelihood group also showed underconnectivity between the right amygdala and the bilateral precuneus, bilateral posterior cingulum, and bilateral occipital regions. Early cerebellar connectivity has been proposed as a regulator of the formation of remote nonmotor networks, with particular significance for autism (Wang, Kloth, & Badura, 2014). Moreover, the early developmental significance of occipital connectivity is also in accordance with studies showing divergent development of binocular rivalry in autism (Robertson, Kravitz, Freyberg, Baron-Cohen, & Baker, 2013), and our findings are in line with past research identifying patterns of underconnectivity in autistic children (Dinstein et al., 2011), in particular underconnectivity between the amygdala and occipital regions (Fishman, Linke, Hau, Carper, & Müller, 2018; Shen et al., 2016). Therefore, it may be that the underpinnings of functional connectivity differences within the brain, found in autistic children, are present *in utero* and during early infancy.

Finally, we found that multiple regions of the brain that showed indications of sex differences were also regions that showed high-autism likelihood-related differences. In fetuses, this effect occurred across multiple brain regions and was most prominent in regions such as the Rolandic operculum, olfactory cortex, gyrus rectus, vermis, precentral gyrus, and amygdala.

In infants, this effect was most prominent in regions such as the medial orbitofrontal cortex, superior occipital gyrus, supplementary motor area, and amygdala. These findings are consistent with previous associations between sexual differentiation and autism (Auyeung et al., 2012; Beacher et al., 2012; Floris et al., 2018; Lai et al., 2013) and, alongside the group-by-sex interaction in fetal total brain volume, further suggest that sexual differentiation and high-autism likelihood differences in brain function and structure may be intertwined.

Regarding the study’s limitations, it is clearly limited by its sample size, particularly by the small size of the high-autism likelihood group and by the attrition of sample size over time, meaning that the results should therefore be considered novel but preliminary. The challenge of collecting viable and comparable longitudinal neuroimaging data from pregnant mothers whose offspring have an increased likelihood for developing autism should not be understated and is likely why an *in utero* study of this design has not previously been conducted. To account for the small sample size, we used the prior literature to direct our focus on total brain volumes and specific seed regions for functional connectivity, as well as applying conservative corrections for multiple comparisons. Even with this cautious approach, we were able to detect findings of altered brain changes over the perinatal period associated with both sex and autism likelihood in brain development, but larger studies are needed to confirm and extend these findings. In addition, none of the infants in this study had received a diagnosis of autism, thus, whilst our findings are relevant to individuals with an increased likelihood for developing autism, they are not necessarily representative of autistic brain development and are therefore not a true test of theories surrounding autism.

In conclusion, these findings indicate neurobiological differences in fetuses with an increased likelihood for developing autism that, with further research, could improve the field’s understanding of the neurobiological basis of autism and eventually aid in the detection of potential neurobiological markers to identify those who may benefit from early screening and interventions. Moreover, we found a link between typical sexual differentiation within the brain and neurobiological differences related to having an increased likelihood for developing autism. This is consistent with the claim that the developmental origins of autism overlap with the development of typical sexual differentiation within the brain (Lai et al., 2013), and may be intertwined.

## Data Availability

Data generated and analyzed during the study is not publicly available due ethical and privacy restrictions.

## Acknowledgements

We are grateful to all the participants for their contribution to this project. We would like to thank the staff at the Rosie Hospital for their contribution to the project, as well as Kate Maxwell for her contribution to the project.

## Declaration of Competing Interest

No authors have any competing interests to declare.

## Funding

This project was funded by the Autism Research Trust. SBC was funded by the Autism Research Trust, the Templeton World Charitable Foundation, and the NIHR Biomedical Research Centre in Cambridge, during the period of this work. This research was funded in whole, or in part, by the Wellcome Trust, award number RG69312, grant number RNAG/528. SBC received funding from the Wellcome Trust 214322\Z\18\Z. For the purpose of Open Access, the author has applied a CC BY public copyright licence to any Author Accepted Manuscript version arising from this submission. Further to this, SBC and RJH also received funding from the Innovative Medicines Initiative 2 Joint Undertaking (JU) under grant agreement No 777394. The JU receives support from the European Union’s Horizon 2020 research and innovation programme and EFPIA and AUTISM SPEAKS, Autistica, SFARI. Disclaimer: The views expressed are those of the author(s) and not necessarily those of the IMI 2JU. The research leading to these results has also received support from the Innovative Medicines Initiative Joint Undertaking under grant agreement n° 115300, resources of which are composed of financial contribution from the European Union’s Seventh Framework Programme (FP7/2007 - 2013) and EFPIA companies’ in-kind contribution. SBC’s research is also supported by the National Institute of Health Research (NIHR) Applied Research Collaboration East of England (ARC EoE) programme. The views expressed are those of the authors, and not necessarily those of the NIHR, NHS or Department of Health and Social Care.

## Notes

### Competing Interest Statement

The authors have declared no competing interest.

### Author Declarations

East of England Cambridge Central REC Committee (REC reference number: 12/EE/0393)

