## supplementary information for "Sex differences in brain development in fetuses and infants who are at low or high likelihood for autism"

***S1)*** *Demographic details of the fetal and infant samples, and their statistical comparisons.*

**Table 1.** Shows demographic details of male and female fetuses in both the typical and high-likelihood groups.

|  | Typical Male  [structural]  (*n* = 15) | Typical Females  [structural]  (*n* =15) | *U* | *p* |
| --- | --- | --- | --- | --- |
| Gestational Age (days) | 221.600  (9.295) | 226.800  (6.784) | 83.000 | 0.219 |
|  | **Typical Males**  **[functional]**  **(*n* = 12)** | **Typical Females**  **[functional]**  **(*n* = 14)** | ***U*** | ***p*** |
| Gestational Age (days) | 227.0833  (6.653) | 221.071  (9.409) | 56.500 | 0.155 |
|  | **High-autism likelihood Males**  **(*n* = 6)** | **High-autism likelihood Females**  **(*n* = 5)** | ***U*** | ***p*** |
| Gestational Age (days) | 224.833  (10.778) | 225.600  (12.219) | 15.000 | 1.00 |

**Table 2.** Shows demographic details of fetuses in the typical and high-likelihood groups.

|  | Typical Group  [structural]  (*n* = 30) | High-autism likelihood Group  [structural]  (*n* = 11) | *t* or χ^2^ | *p* |
| --- | --- | --- | --- | --- |
| Gestational Age (Days) | 224.200  (8.422) | 225.182  (10.861) | 0.306 | 0.761 |
| Sex (% Female) | 50.0 | 45.5 | 0.067 | 0.796 |
|  | **Typical Group**  **[functional]**  **(*n* = 26)** | **High-autism likelihood Group [functional]**  **(*n* = 11)** | ***t* or χ^2^** | ***p*** |
| Gestational Age (Days) | 223.846  (8.652) | 225.182  (10.861) | 0.398 | 0.693 |
| Sex (% Female) | 53.8 | 45.5 | 0.218 | 0.641 |

**Table 3.** Shows demographic details of male and female infants in both the typical and high-likelihood groups.

|  | Typical Male  [structural]  (*n* = 10) | Typical Females  [structural]  (*n* = 10) | *t* | *p* |
| --- | --- | --- | --- | --- |
| Age Since Conception (days) | 354.000  (13.548) | 354.100  (13.304) | 0.0167 | 0.987 |
| Birthweight (grams) | 3564.000  (462.594) | 3265.500  (363.986) | 1.604 | 0.126 |
|  | **Typical Males**  **[functional]**  **(*n* = 9)** | **Typical Females**  **[functional]**  **(*n* = 7)** | ***t*** | ***p*** |
| Age Since Conception (days) | 353.778  (14.351) | 354.571  (15.608) | 0.106 | 0.917 |
| Birthweight (grams) | 3526.667  (474.408) | 3265.000  (398.560) | 1.171 | 0.261 |
|  | **High-autism likelihood Males**  **(*n* = 4)** | **High-autism likelihood Females**  **(*n* = 3)** | ***t*** | ***p*** |
| Age Since Conception (days) | 357.250  (13.301) | 348.333  (24.583) | 0.626 | 0.559 |
| Birthweight (grams) | 4025.750  (189.328) | 3558.667  (596.022) | 1.512 | 0.191 |

**Table 4.** Shows demographic details of infants in the typical and high-likelihood groups.

|  | Typical Group  [structural]  (*n* = 20) | High-autism likelihood Group  [structural]  (*n* = 7) | *t* or χ^2^ | *p* |
| --- | --- | --- | --- | --- |
| Age Since Conception (days) | 354.050  (13.067) | 353.429  (17.681) | 0.393 | 0.698 |
| Sex (% Female) | 50.0 | 42.9 | 0.106 | 0.745 |
| Birthweight (grams) | 3414.750  (433.093) | 3825.571  (445.724) | 2.162 | 0.040 |
|  | **Typical Group**  **[functional]**  **(*n* = 16)** | **High-autism likelihood Group**  **[functional]**  **(*n* = 7)** | ***t* or χ^2^** | ***p*** |
| Age Since Conception (days) | 354.125  (14.403) | 353.429  (17.681) | 0.010 | 0.9215 |
| Sex (% Female) | 43.8 | 42.9 | 0.002 | 0.968 |
| Birthweight (grams) | 3412.188  (448.939) | 3825.571  (445.724) | 2.036 | 0.055 |

***S2)*** *Non-significant results.*

Fetal Brain Structure

There were no sex differences in total brain volume within the typical group, *F*(1, 27) = 1.752, *p* = 0.197. There were also no differences between the typical and high-autism likelihood fetuses in total brain volume, *F*(1, 37) = 0.213, *p* = 0.647.

Fetal Brain Function

There were no sex differences in resting-state functional connectivity within the typical group. There were no significant group-by-sex interactions in resting-state functional connectivity.

Infant Brain Structure

There was no sex difference in total brain volume within the typical group, *F*(1, 16) = 2.741, *p* = 0.117, and there were no differences in total brain volume between the high-autism likelihood and typical groups, *F*(1, 22) = 3.366, *p* = 0.080. There was no longer a group-by-sex interaction in total brain volume in 3 month old infants, *F*(1, 21) = 0.544, *p* = 0.469; **Figure 1**.

**Figure 1.** Shows infant total brain volumes, where there was no longer a significant group-by-sex interaction.

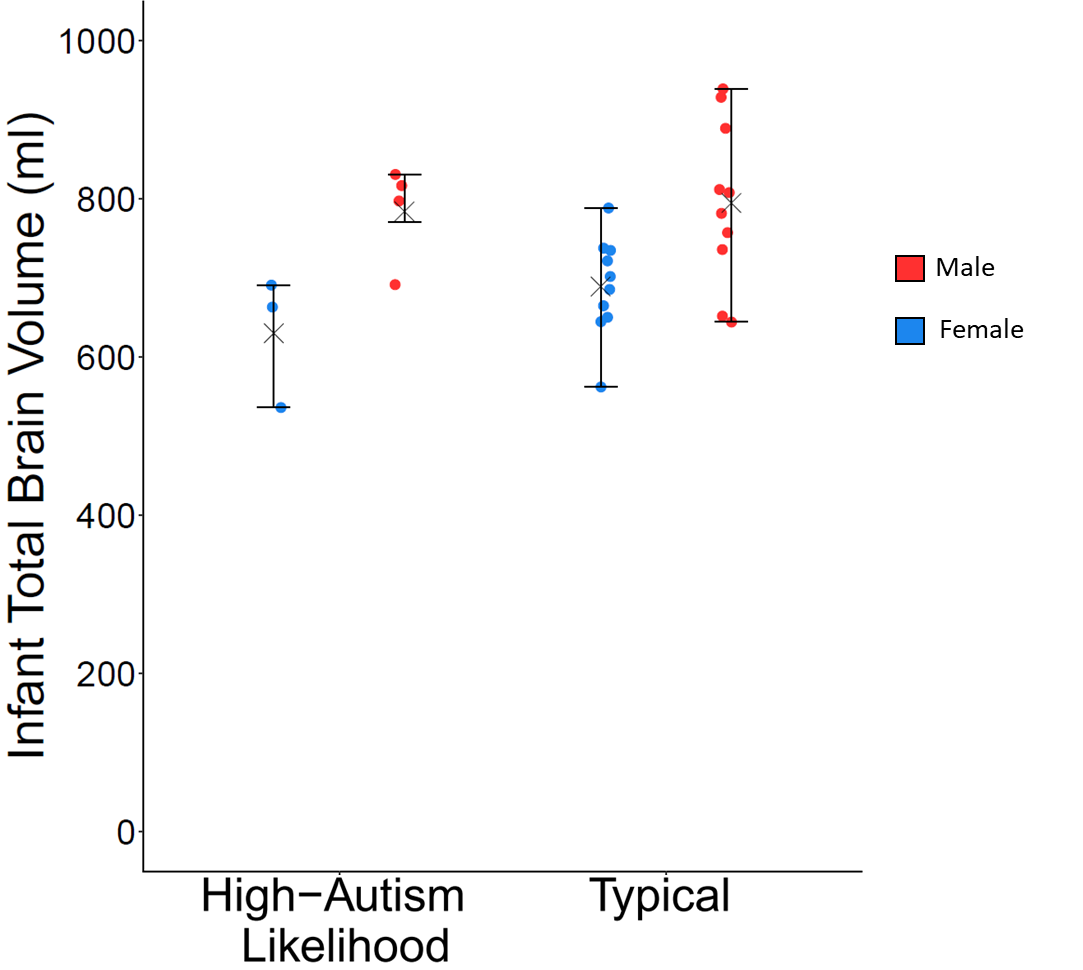

Infant Brain Function

There were no sex differences within the typical infant group in resting-state functional connectivity. There were no group-by-sex interactions in resting-state functional connectivity.

Interaction Effects with Time

There was no significant group-by-time interaction in total brain volume, *F*(1, 21) = 1.518, *p* = 0.232, nor were there significant sex-by-time or group-by-time interactions in resting-state functional connectivity.

***S3)*** *Further details of the correlations, shown as r^2^, between sex-difference and group difference F-scores, for all six seed regions.*

The fetal *r*^2^ values of positive correlations ranged between 0.0108 - 0.4146, with the median *r*^2^= 0.0544 and mean *r*^2^= 0.0871. The fetal *r*^2^ values of negative correlations ranged between 0.0133 - 0.4391, with the median *r*^2^= 0.0606 and mean *r*^2^= 0.0835. The frequency distributions of positive and negative fetal correlations are shown in **Figure 2**.

The infant *r*^2^ values of positive correlations ranged between 0.0136 - 0.3562, with the median *r*^2^= 0.0725 and mean *r*^2^= 0.0920. The infant *r*^2^ values of negative correlations ranged between 0.0129 - 0.1837, with the median *r*^2^= 0.0443 and mean *r*^2^= 0.0573. The frequency distributions of positive and negative infant correlations are shown in **Figure 3**.

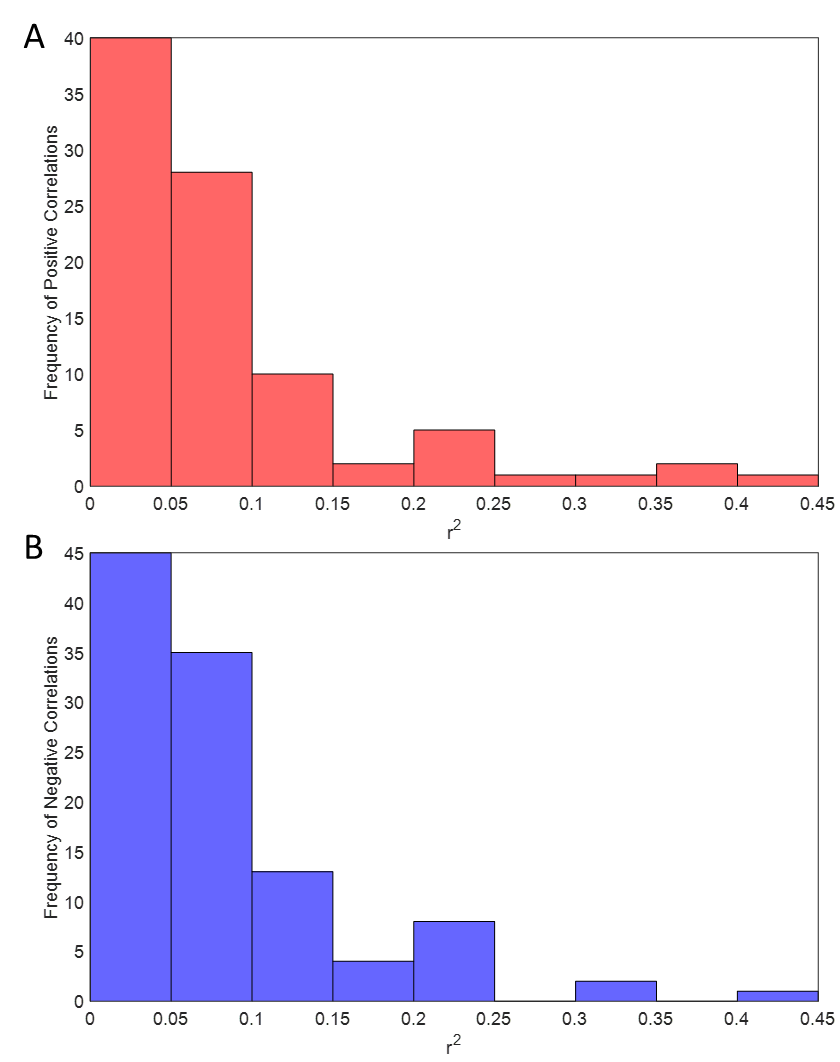

**Figure 2.** Shows the frequency distribution of positive correlations (A) and negative correlations (B) at the fetal timepoint.

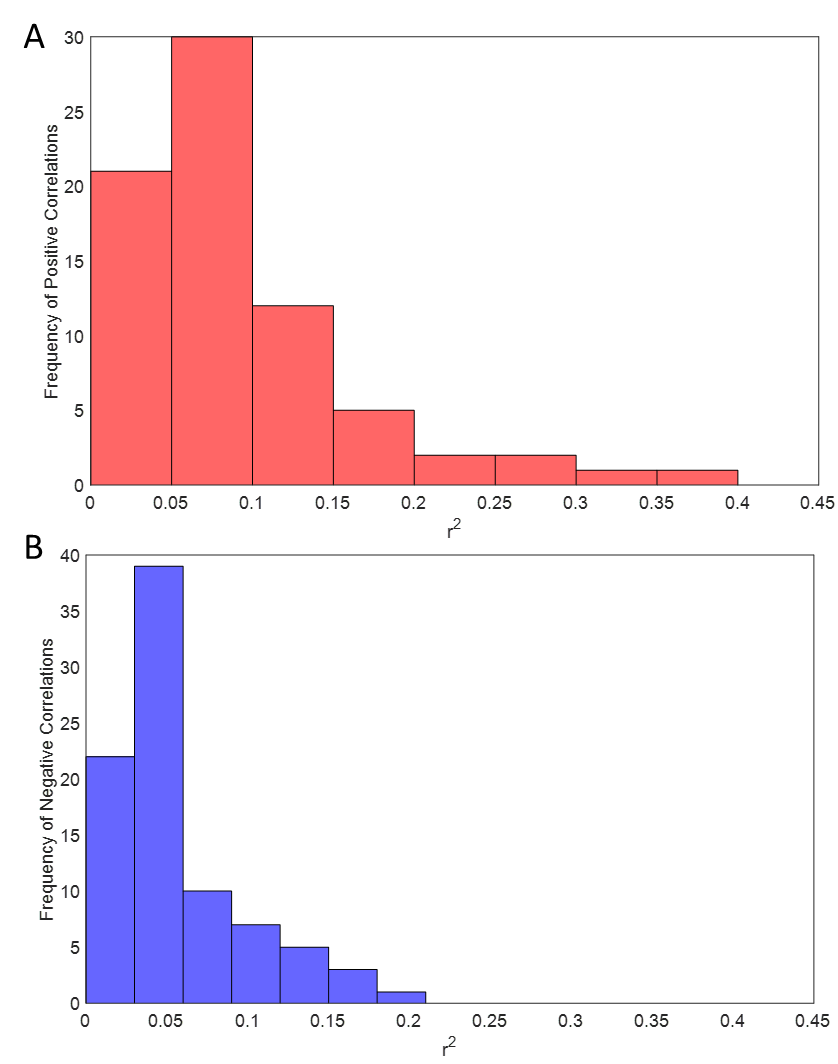

**Figure 3.** Shows the frequency distribution of positive correlations (A) and negative correlations (B) at the infant timepoint.

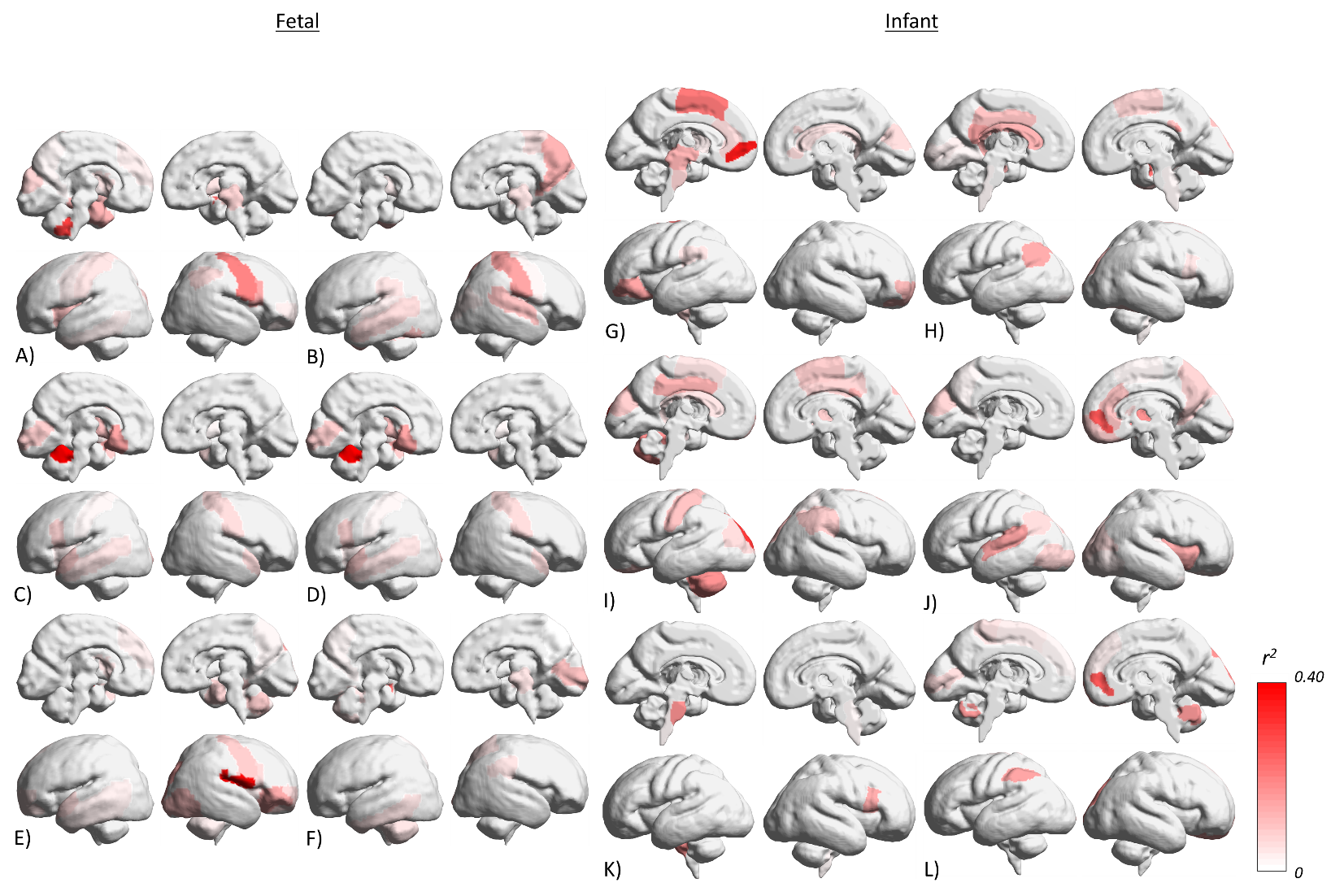

**Figure 4.** Shows the r^2^ values across the six seed regions for both fetuses and infants seed, when correlating sex difference effects with autism high-likelihood-related differences.

**Fetal**: A) Left Amygdala Seed, B) Right Amygdala Seed, C) Left Hippocampus Seed, D) Right	Hippocampus Seed, E), Left Cerebellum Seed, F) Right Cerebellum Seed.

**Infant**: G) Left Amygdala Seed, H) Right Amygdala Seed, I) Left Hippocampus Seed, J) Right	Hippocampus Seed, K), Left Cerebellum Seed, L) Right Cerebellum Seed.

**Table 5.** Shows the fetal seed regions’ correlations between sex difference and group difference *F* scores, in order of effect size, with *r*^2^ in parentheses.

| Seed Regions | | | | | |
| --- | --- | --- | --- | --- | --- |
| Left Amygdala | **Right Amygdala** | **Left Hippocampus** | **Right Hippocampus** | **Left Cerebellum** | **Right Cerebellum** |
| Central Vermis Left  (0.294) | Postcentral Gyrus Right  (0.149) | Anterior Vermis Left  (0.415) | Rectus Left  (0.357) | Rolandic Operculum Right  (0.383) | Amygdala Left  (0.226) |
| Amygdala Right  (0.232) | Precuneus Right  (0.117) | Anterior Vermis Right  (0.231) | Inferior Frontal Gyrus Left  (0.328) | Orbital Part of Inferior Frontal Gyrus Right  (0.136) | Calcarine Sulcus Right  (0.098) |
| Precentral Gyrus Right  (0.199) | Fusiform Gyrus Left  (0.098) | Olfactory Cortex Left  (0.143) | Paracentral Lobule Right  (0.226) | Superior Occipital Gyrus Right  (0.131) | Midbrain Right  (0.049) |
| Inferior Frontal Gyrus Right  (0.126) | Superior Temporal Right  (0.096) | Putamen Left  (0.088 | Central Vermis Left  (0.221) | Inferior Occipital Cortex Right  (0.094) | Cerebellum Left  (0.048) |
| Parahippocampus Left  (0.114) | Middle Temporal Gyrus Left  (0.060) | Superior Temporal Pole Left  (0.084) | Olfactory Cortex Left  (0.156) | Posterior Vermis Right  (0.085) | Inferior Temporal Gyrus Left  (0.047) |
| Insula Left  (0.093) | Paracentral Lobule Right  (0.050) | Calcarine Sulcus Left  (0.084) | Rectus Right  (0.117) | Precentral Gyrus Right  (0.083) | Superior Parietal Lobule Right  (0.042) |
| Midbrain Right  (0.072) | Midbrain Right  (0.050) | Inferior Frontal Gyrus Left  (0.064) | Superior Frontal Gyrus Left  (0.107) | Parahippocampus Right  (0.068) | Supramarginal Gyrus Right  (0.039) |
| Cuneus Left  (0.066) | Supramarginal Gyrus Left  (0.045) | Superior Temporal Pole Right  (0.061 | Cuneus Right  (0.084) | Superior Temporal Pole Left  (0.059) | Precuneus Left  (0.017) |
| Inferior Parietal Lobule Right  (0.052) | Rolandic Operculum Right  (0.036) | Postcentral Gyrus Right  (0.058) | Middle Frontal Gyrus Right  (0.073) | Inferior Frontal Gyrus Right  (0.053) | Precuneus Right  (0.011) |
| Caudate Left  (0.050) | Middle Occipital Gyrus Right  (0.028) | Middle Temporal Gyrus Left  (0.039) | Orbital Part of Inferior Frontal Gyrus Left  (0.064) | Orbital Part of Middle Frontal Right  (0.050) |  |
| Postcentral Gyrus Left  (0.048) | Superior Temporal Gyrus Left  (0.027) | Insula Left  (0.029) | Supplementary Motor Area Left  (0.062) | Orbital part of Middle Frontal Gyrus Left  (0.049) |  |
| Putamen Right  (0.041) | Putamen Left  (0.027) | Putamen Right  (0.025) | Midbrain Right  (0.056) | Caudate Left  (0.041) |  |
| Medial Frontal Gyrus Left  (0.035) | Precentral Gyrus Right  (0.021) | Postcentral Gyrus Left  (0.023) | Anterior Cingulate Left  (0.052) | Insula Right  (0.035) |  |
| Orbital Part of Middle Frontal Right  (0.033) |  |  | Medial Frontal Gyrus Left  (0.050) | Inferior Temporal Gyrus Left  (0.030) |  |
| Putamen Left  (0.032) |  |  | Thalamus Right  (0.022) | Medial Frontal Gyrus Left  (0.030) |  |
| Inferior Temporal Gyrus Left  (0.029) |  |  | Precentral Gyrus Right  (0.018) | Middle Temporal Gyrus Left  (0.028) |  |
| Precentral Gyrus Left  (0.029) |  |  |  | Cerebellum Right  (0.027) |  |
| Rolandic Operculum Left  (0.021) |  |  |  | Precuneus Right  (0.019) |  |
| Precuneus Left  (0.014) |  |  |  |  |  |

**Table 6.** Shows the infant seed regions’ correlations between sex difference and group difference *F* scores, in order of effect size, with *r*^2^ in parentheses.

| Seed Regions | | | | | |
| --- | --- | --- | --- | --- | --- |
| Left Amygdala | **Right Amygdala** | **Left Hippocampus** | **Right Hippocampus** | **Left Cerebellum** | **Right Cerebellum** |
| Medial Orbitofrontal Gyrus Left  (0.356) | Amygdala Right  (0.223) | Superior Occipital Gyrus Left  (0.312) | Medial Orbitofrontal Gyrus Right  (0.255) | Medulla Left  (0.166) | Medial Orbitofrontal Gyrus Right  (0.256) |
| Supplementary Motor Area Left  (0.209) | Posterior Cingulate Left  (0.143) | Anterior Vermis Left  (0.169) | Putamen Right  (0.166) | Inferior Frontal Gyrus Right  (0.135) | Superior Occipital Gyrus Right  (0.163) |
| Orbital Part of Inferior Frontal Gyrus Left  (0.146) | Angular Gyrus Left  (0.134) | Midcingulate Left  (0.119) | Superior Temporal Pole Left  (0.146) | Cerebellum Right  (0.020) | Central Vermis Right  (0.156) |
| Pons Left  (0.118) | Midbrain Left (0.118) | Postcentral Gyrus Left  (0.098) | Insula Right  (0.126) |  | Inferior Parietal Lobule Left  (0.140) |
| Orbital Part of Middle Frontal Right  (0.109) | Midcingulate Left  (0.099) | Superior Occipital Gyrus Right  (0.091) | Thalamus Left  (0.095) |  | Orbital part of Superior Frontal Gyrus Right  (0.125) |
| Medulla Left  (0.064) | Superior Occipital Gyrus Right  (0.092) | Inferior Parietal Lobule Right  (0.084) | Anterior Cingulate Right  (0.092) |  | Calcarine Sulcus Left  (0.060) |
| Anterior Cingulate Left  (0.051) | Supplementary Motor Area Right  (0.067) | Putamen Right  (0.082) | Rectus Right  (0.079) |  | Paracentral Lobule Left  (0.038) |
| Cuneus Right  (0.050) | Fusiform Gyrus Right  (0.064) | Orbital part of Superior Frontal Gyrus Left  (0.081) | Inferior Occipital Cortex Left  (0.069) |  | Supplementary Motor Area Left  (0.031) |
| Midbrain Right  (0.041) | Inferior Frontal Gyrus Right  (0.042) | Cuneus Left  (0.081) | Cuneus Left  (0.064) |  | Medial Frontal Gyrus Right  (0.028) |
| Supramarginal Gyrus Left  (0.041) | Calcarine Sulcus Left  (0.029) | Supplementary Motor Area Right  (0.078) | Precuneus Right  (0.063) |  | Medial Frontal Gyrus Left  (0.025) |
| Putamen Left  (0.038) | Lingual Gyrus Left  (0.024) | Supramarginal Gyrus Right  (0.076) | Middle Occipital Gyrus Right  (0.059) |  |  |
|  | Cerebellum Right  (0.024) | Midcingulate Right  (0.065) | Angular Gyrus Left  (0.057) |  |  |
|  |  | Middle Occipital Gyrus Left  (0.063) | Caudate Right  (0.054) |  |  |
|  |  | Midbrain Left  (0.039) | Superior Occipital Gyrus Right  (0.051) |  |  |
|  |  | Supplementary Motor Area Left  (0.039) | Orbital part of Superior Frontal Gyrus Right  (0.042) |  |  |
|  |  | Superior Parietal Lobule Right  (0.021) | Inferior Occipital Cortex Right  (0.035) |  |  |
|  |  |  | Supplementary Motor Area Right  (0.023) |  |  |
|  |  |  | Superior Temporal Right  (0.021) |  |  |
|  |  |  | Inferior Temporal Gyrus Right  (0.020) |  |  |
|  |  |  | Precuneus Left  (0.014) |  |  |
